# Understanding community leaders’ and program coordinators’ perceptions of and needs from a community systems map: a qualitative study

**DOI:** 10.1101/2024.01.26.24301839

**Authors:** Margret Lo, Natasha Ross, Sonia S. Anand, Russell J de Souza, Gita Wahi, Dipika Desai, Sujane Kandasamy, Diana Sherifali, Deborah DiLiberto, K. Bruce Newbold, Mary Crea-Arsenio, Patty Montague, Fatimah Jackson-Best

## Abstract

**Objectives:** A systems map for the Riverdale community in Hamilton, Ontario has documented programs, activities, and spaces for healthy, nature-based activities. A systems map was created to provide an approachable resource for community members. As a means of understanding whether this map would prove useful, interviews with community and organization leaders were conducted. The objectives of this study were to 1) characterize user engagement with recreational and active living programs and services located in and around the Riverdale community, 2) determine existing barriers to and facilitators of accessing recreational and active living programs for those in the Riverdale community, as perceived by those who deliver programs and, 3) improve the functionality and usability of the system map, including any information or services that may be missing.□□

**Methods:** Twenty 60-minute semi-structured interviews were conducted with community leaders and program coordinators who focus on healthy active living and work within the Riverdale community.

**Results:** We learned that Riverdale programming is typically gender-specific, women-dominated, and targeted to children and youth. Additionally, cost, transportation and accessibility, and language were the greatest barriers to accessing recreational programming. Conversely, facilitators for program involvement included transportation assistance, compensation, and food provision. Finally, interviews revealed the need for sustainability, clarity, and language options within the systems map.

**Conclusion:** This work procured the necessary information to edit the interactive systems map according to community needs. With this map, Riverdale community members are able to access a source of information about activities and services.

## Introduction

In 2021, Canada welcomed more than 401,000 immigrants (Government of Canada, 2021). Although Canada provides a host of benefits like multiculturalism and stability, newcomers often face many challenges such as financial barriers, attaining and maintaining optimal health and wellness, stable employment, and house ownership (Weitz, 2011; Lane et al., 2021). Recent research has indicated that immigrants, mostly those of visible minorities develop diabetes earlier compared to their white Caucasian counterparts (Tenkorang, 2017). Further, certain ethnic groups are at a higher risk of developing obesity, hypertension, and poor mental health outcomes (Wahi et al., 2014; Sahoo et al., 2015).

In Hamilton, Ontario, the Riverdale neighbourhood serves a large proportion of visible minorities (City of Hamilton, 2021; CBC News, 2017). Compared to other areas of Hamilton, Riverdale experiences higher material deprivation. The Strengthening Community Roots: Anchoring Newcomers in Wellness & Sustainability (SCORE!, https://okanagan.mcmaster.ca/initiatives/score/) project aims to co-design an intervention that nurtures and optimizes healthy active living to prevent obesity and related cardio-metabolic risk factors among newcomer children and families in Hamilton (Wahi et al., 2023).

The SCORE! systems map was created to help reduce barriers for local families in accessing local recreation services. “Systems mapping” is an approach that is used to identify and present aspects of a system in a comprehensive manner, as a means of making complex systems seem approachable (Canadian Center on Substance Abuse, 2014). The SCORE! systems map was created using a typical systems mapping approach. With the help of community members, key spaces in Hamilton, Ontario that offer programs, activities, and spaces for healthy nature-based activities were documented. To understand the potential, the impact, and the utility of the system map for the residents in Riverdale, qualitative interviews were conducted. This work aims to accomplish three main objectives, 1) characterize user engagement with recreational and active living programs and services located in and around the Riverdale community; 2) determine existing barriers and facilitators to accessing recreational and active living programs experienced by residents in the Riverdale community, as perceived by community leaders and organizations; and 3) improve the functionality and usability of the system map, including any information or services that may be missing.

### Ethics Approval

This study was approved by the Hamilton Integrated Research Ethics Board (HIREB #15120). Participants were asked to provide informed consent upon participating.

## Method

### Design

The SCORE! project takes in community-based participatory research (CBPR) approach. This approach encourages the participation of community members in its research process and helps to ensure that research is being done in a manner that focuses on communities’ needs (Flicker et al., 2007). In doing so, interventions are often more likely to be accepted by community members and can be better suited for providing support (O’Brien & Whitaker, 2011).

### Setting

Riverdale is a neighbourhood located in Hamilton, Ontario, Canada where 50% of the residents were born outside of Canada, and 16% are recent immigrants (Statistics Canada, 2021). This percentage intersects with a quarter (25%) of families immigrating to Canada in the last 20 years, and 26% identifying as low income as per Statistics Canada’s low-income measures (Census Mapper, 2021; Statistics Canada, 2021). As a city, 7% of Hamilton’s immigrant population is located in Riverdale – making it home to the highest proportions of recent immigrants in the city (Statistics Canada, 2016).

### Recruitment

Individuals were included in this study if they were 18 years of age or older and were able to conduct an interview in English. Further, individuals had to be considered a community organizer, community advocate, or community leader for an organization that serves the multi-lingual, multi-cultural, and multi-ethnic community of Riverdale or the City of Hamilton. Participants were recruited using snowball sampling. The SCORE! project works closely with Riverdale organizations and collaborated with these organizations to get recommendations and referrals from other organizations of interest to the project. At the end of each interview, participants were asked to suggest fellow community leaders who may be important to contact given the context of the interview.

### Interviews

To assess the community’s needs, qualitative interviews were conducted to discuss the systems map and elicit perspectives regarding what services should be reflected within the map. Between February and May 2023, 20 semi-structured interviews were conducted with community leaders and program coordinators over Zoom by one of the study’s authors (ML). Participants provided informed consent prior to the beginning of each interview. Upon completion of the interview, participants received a $25 gift card that was sent via an online platform.

Questions were asked to explore three broad categories: 1) organization and program information of services, 2) user engagement in healthy active living programming, 3) systems map feedback. These questions were open-ended and allowed for community leaders and program coordinators to openly discuss their experiences. Through these questions, researchers could characterize user engagement in healthy active living programming, determine existing barriers and facilitators to accessing these services, and improve the functionality and usability of the systems map. Appendix A contains the interview guide that was used during these qualitative interviews.

### Analysis

Interviews were audio recorded over Zoom and transcribed by one of the authors (ML) with the assistance of Otter.ai, a transcription software. Upon completion of transcription, one of the authors (ML) used NVivo (Lumivero, United States of America) to code data according to the six steps for conducting thematic analysis as outlined by Braun and Clark (2006). A codebook was created to monitor all codes and themes. Data was organized into codes which were then organized into larger themes by a single researcher. These themes reflected the main objectives of this study. After coding information, the themes were reviewed for relevance and organization. Similar themes were combined. For this report, themes that were considered relevant where those that had at least three references from the interviews.

### Positionality Statement

Prior to presenting these findings, the primary author would like to acknowledge her positionality. The primary author led the data collection and analysis processes. She brings her perspectives as a female, East Asian/Southeast Asian, settler, cis-gendered, Master of Public health graduate in Canada. As a child of an immigrant and a refugee, she was able to develop a rapport with study participants. While the primary author strived to be reflexive on position, privilege, and perspective in data collection and data analysis, her perspective may influence the results and interpretation. To avoid speaking for this data, the primary author engaged in reflexive processes throughout the entirety of this study. To do so, the author worked with FJB to ensure the study was guided by shared expertise. Regular team meetings were held to ensure that the study remained relevant, and appropriate to the community context.

## Results

### Participant Characteristics

We recruited 20 individuals, all of which were recruited through snowball sampling. Interviews ranged from 36 to 74 minutes and lasted an average of 46 minutes. Participants described their organization’s programs and services, the types of users that engaged with their programs, and whether or not they saw the systems map as being a useful tool for those in the local Riverdale community.

### Recreational and Active Living Program User Engagement Characteristics

Participants in these qualitative interviews, discussed the types of users that have engaged in their healthy active living programming (Figure 1).

**Fig 1.**
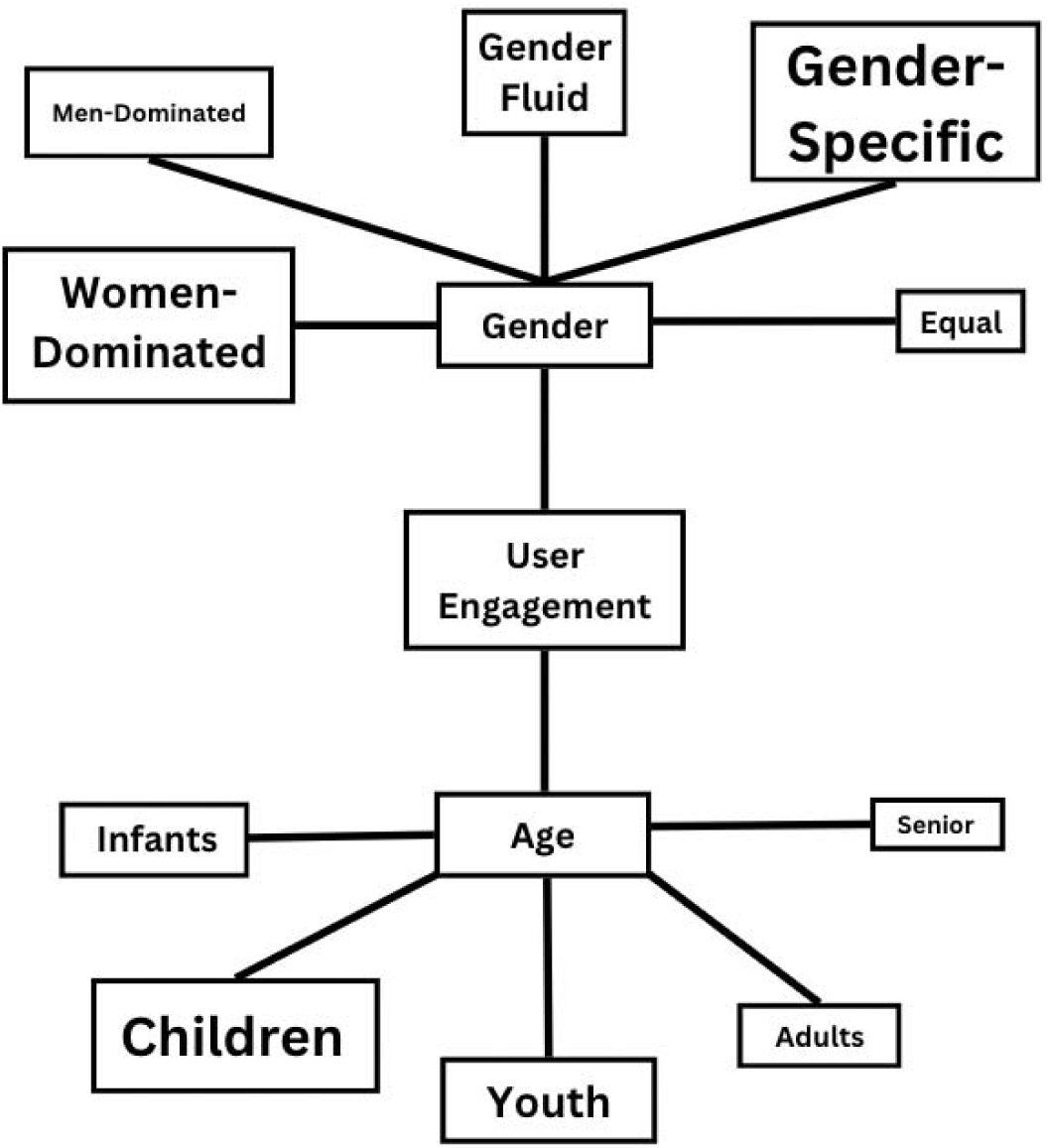
Themes associated with user engagement in active living programs. *Size of themes indicate their prevalence in analysis. Larger items indicate greater frequency in discussion.

#### Gender

Interviewees consistently mentioned the role that gender played in user engagement. For these community leaders, gender and their respective roles within the Riverdale community were significant considerations that these leaders made when planning and organizing healthy active living programming. Despite there being programming available for women and men, programs were often gender-specific, or women-dominated.

Similarly, program leaders were often influenced by the community’s needs. One community leader mentioned that “[they]’ve been visioning and listening to the community and hearing all the gaps”. Another leader mentioned that they “adapt some services… according to the needs”. One of these needs focused on the importance of gender-specific programs. Several organizations highlighted the processes that they took when recognizing the needs of the Riverdale community.

“We noticed that through the pandemic, there [were] a lot of family issues, violence among the families. And so, we created the women gardening group, as soon as the government gave us that space.” Although gender-specific programs are needed within the Riverdale community, few programs discussed options for those exploring their sexual and gender identity. Community partners have addressed that “there are certainly children who are exploring and, you know, determining, and you know, figuring out, they’re exploring their gender identity”. However, a few community partners also discussed that the last time an individual had self-identified as gender fluid was “maybe five, or six years ago”.

#### Age

The age of children were significant factors of the type of programming that could be delivered in the Riverdale community. Many community leaders identified that programming has been developed to “have…all ages”, indicating that many Riverdale programs are developed to serve all populations in Riverdale. Despite programming being offered to all ages within the Riverdale community, awareness of such programming to the intended populations remains unknown. It is clear, however, that the majority of programming has been made with children, youth, and infants in mind. As a result, there has been less of a focus on programming for both adults and senior citizens.

### Barriers to Accessing Recreational and Active Living Programs

When talking about the programming and recreational activities that local community partners offer, interviewees were eager to talk about the barriers associated with accessing these services (Figure 2). Given Riverdale’s demographic composition, it is understood by organizational leaders that there are several barriers when accessing programs, this includes financial barriers. When discussing the barriers that community members faced when accessing specific healthy active living programs, barriers were often related to this financial stress.

**Fig 2.**
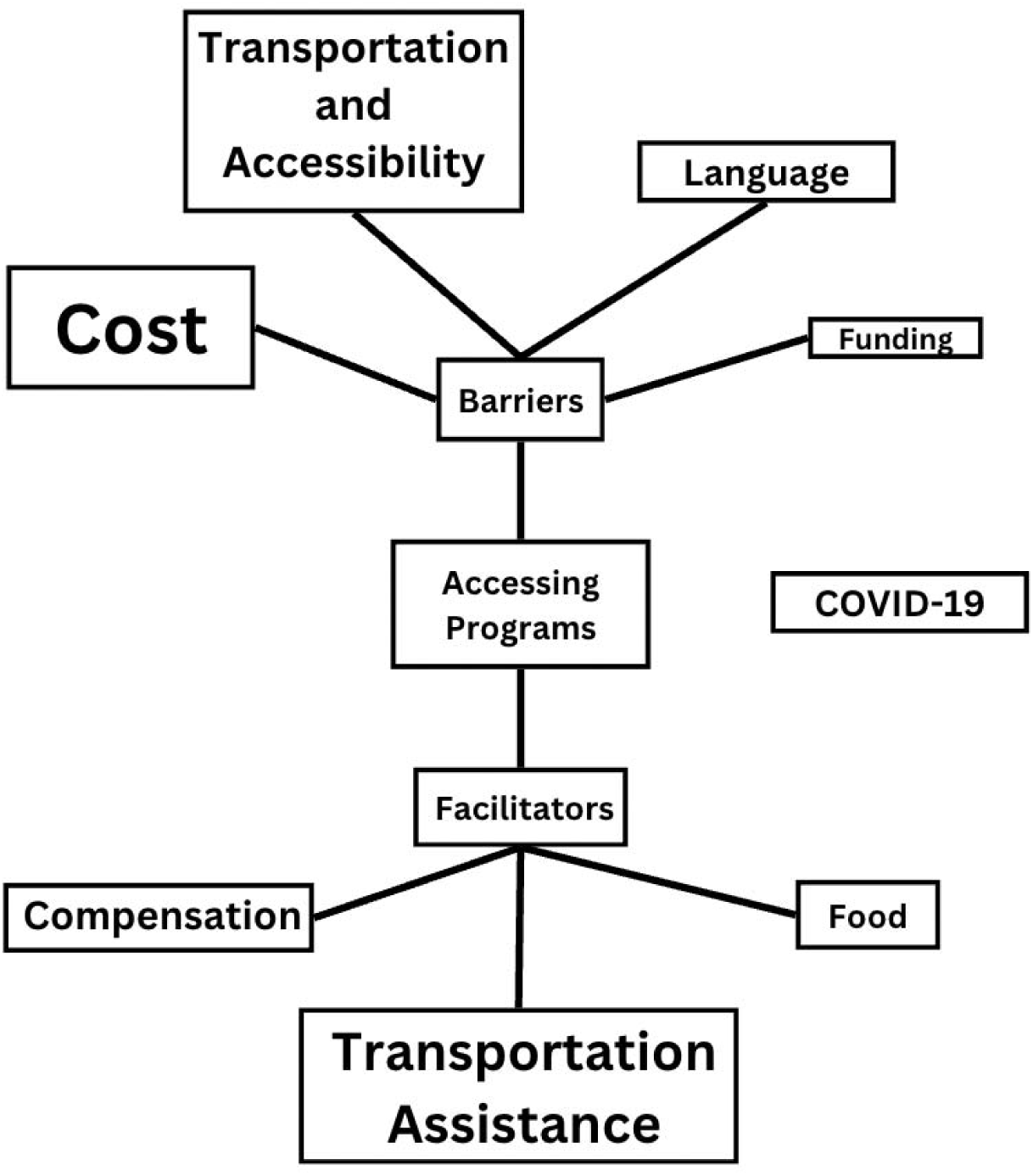
Themes associated with accessing programs in the Riverdale community. *Size of themes indicate their prevalence in analysis. Larger items indicate greater frequency in discussion.

#### Cost

Community leaders often recognized that the cost of programs was a major barrier for Riverdale residents. Despite some subsidy programs to alleviate these costs, accessing these programs was still difficult for members of the Riverdale community due to stigma of financial instability. The quote below further contextualizes this issue.

> I mean, cost is occasionally the thing that pops up quite often. You know, we offer a fee assistance program. But that doesn’t always mean that, you know, the kids take the forms home to their parents or that their parents are willing to fill it out. There you know, there’s some stigma around having to admit how much or how little money you make, and then putting it on a document for someone to assist you with [is difficult].

Through discussion with community leaders, it is clear that the cost of programs, and accepting help for such costs, were difficult for most individuals – speaking to the confluence of stigma, economic barriers, and issues surrounding anonymity.

#### Transportation and Accessibility

Apart from the barriers associated with the cost of programs, transportation and accessibility proved to be another significant barrier for accessing local activities. In fact, one interviewee noted, “the barriers, huge barriers are transportation, always transportation”. As a result of a lack of transportation, community members are not able to attend as many healthy active living and community-building events.

> So, transportation is actually a big thing. A lot of people mentioned that, you know, you’re not able to come to a lot of the programs because of transportation. Transportation is an issue.

Transportation to and from community partner locations posed a concern. But in a similar vein, transportation within the community partner locations were also points of contention for many.

> [Our organization] itself is next to the creek and so the water table is high. And so, oftentimes, like…when it’s raining, it gets really, really muddy. And so, folks who may be using a wheelchair or some mobility device, they really do have a hard time accessing the [organization] because there’s nothing to like grasp to access [it]. And so that really like poses a huge challenge for them to be able to get into the [organization] comfortably and moving around [it] comfortably because there also [aren]’t like accessible pathways.

Regardless of whether events were held indoors or outdoors, access to these events, and access within these events have proven to be a barrier for community participation. Accessibility of events has been a large area of concern, highlighting its importance in promoting community participation.

#### Language

Many program coordinators and community leaders indicated the community’s need for a sense of belonging. Despite many efforts to promote this sense of community, a barrier for many of these programs are the language barriers that exist between program staff and program participants.

> So thankfully, [our organization has] a lot of different staff who speak a lot of different languages who have supported our placement office. Our placement office is the first point of contact for families… And translation is a part of that as well. So, if we cannot find… community [translators] to support them through that process, I would say probably the language… is a barrier for folks.

As mentioned, in an attempt to mitigate barriers, many individuals have staff on hand who speak a variety of languages. Consequently, whether organizations can help to navigate language barriers, depends on whether organizations have the appropriate staff. For instance, in another organization the situation was markedly different, and an interviewee stated, “if we have instructors that can teach [in that language] … then we can offer [those programs] but, yeah, we don’t have a lot of different languages being spoken in our facilities”.

#### Funding

Apart from costs associated with accessing programs, program coordinators also identified that lack of funding is a major issue for running and organizing programming. This lack of funding means that organizations are “pretty bound with what [they]’re able to do with [their] funding”, and that organizations are “limited”. Ultimately, a lack of funding indicates a restriction of programming, whether that restriction is specific to the audience programming is provided to, as well as the timelines and frequency in which programming can happen. As a result, this often results in individuals being excluded from programming that they may want and need to participate in.

### Facilitators to Accessing Recreational and Active Living Programs

Community leaders were also eager to discuss facilitators to program participation (Figure 2). When discussing facilitators associated with accessing healthy active living programming, these almost always directly paralleled the barriers that were discussed (Table 1). Local organizations and community leaders are aware of the barriers to their programs and have done work to mitigate these.

**Table 1.** Paralleling barriers and facilitators associated with healthy active living programming in Riverdale.

| **Barrier** | **Facilitator** |
| --- | --- |
| Cost | Compensation<br>Food |
| Transportation and Accessibility | Transportation Assistance |
| Language |  |
| Funding |  |
| COVID-19 |  |

#### Transportation Assistance

The barrier of transportation has called community partners to focus on a solution to their organization’s lack of accessibility. As previously discussed, transportation has been an obstacle for many individuals when it comes to navigating and participating in programs. In interviews, many interviewees discussed the ability to provide bus tickets for families. One organization discussed how “[they] can provide bus tickets for supporting families and … can help them access free [transit] cards for children and whatnot”. In addition to providing transportation support to program attendees, organizations have also provided these means of transportation for their volunteers – who were identified as essential unpaid staff members who lead the healthy active living programming for the Riverdale community. One organization mentioned that they “do give bus tickets, for example, to compensate the time for the volunteers who work with us”. For situations in which a more rapid form of transportation is needed, community leaders have also discussed the ability to provide other forms of transportation. However, due to a lack of funding, these forms of transportation have proven more difficult to acquire. One community partner discussed that “[they] will provide taxi chits…if it’s an emergency”. Transportation is a significant factor that impairs or promotes the ability of local community members to attend events. Consequently, many community leaders have turned to supplying some form of transportation so that Riverdale community members can actively attend healthy active living events and programming.

#### Compensation

One barrier that was mentioned was the cost of participation in programs. As a result, community partners have identified compensation as a method of incentivizing and facilitating participation in local programming. Despite a lack of funding, community partners expressed the desire and need to compensate individuals for their time and participation as much as possible.

> [We have] very limited resources for compensation. But I partner with others and try to find other resources to compensate our clients when they participate in activity, either with us or with our agency.

In particular, programming targeted at youth and children often offered some form of compensation. Community leaders recognized that when children wanted to attend programming, their parents often tried to ensure that their children could at least attend one session. One community leader recognized that “[they] give prizes sometimes, like we have contests and stuff like that … the prizes that were given were like hoverboards and drones, and stuff like that”. Overall, community partners discussed how they try to reduce the barriers that individuals often face when accessing programs by being a “one stop shop” that offers a variety of services, which lessened participants going between multiple sites, fostering increased efficiency.

#### Food

With the Riverdale community largely serving a significant newcomer population, the incurred cost of coming to healthy active living programming often competes with daily living costs, such as food. Consequently, many organizations have opted to provide meals as a way to ensure that individuals feel as if attending recreational programs does not impair their overall quality of life.

> We don’t have a lot of barriers. Let’s say the biggest barriers are age, transportation, food. So, we’re kind of providing those things, transportation, food, increasing our age, who we can serve.

Often, food served as a way to incentivize individuals, so organizations could take the time to educate individuals about their services and supports. One organization mentioned the creation of a “family dinner night…that will be once a week, and again we’ll have a team of people that can help support families”. In particular, food was often mentioned as a source of connection for individuals. Food provided a sense of community. Even when individuals were not able to attend programming, food was always a way of reinforcing connection.

> If they’re not there for some reason, they’re sick, then the food that we have, we make sure we put some food on the side and [whoever lives in that area] … they will drop off the food. Then, we get phone calls of thank yous and how they appreciate it.

Overall, food proved to be a large facilitator for enticing community participation, while also maintaining participation and connectedness.

### COVID-19

The COVID-19 pandemic was also mentioned as both a barrier and facilitator for many programs in the Riverdale neighbourhood. Throughout the pandemic, some organizations expressed that their services were more greatly sought after. In fact, “during COVID times we got more and more calls”. Some organizations saw a greater need for their services and have been able to maintain the relationships that they developed through the pandemic. In a similar vein, organizations saw that COVID-19 acted as a way for innovation to take place. Many community organizations were forced to “shift … from [developing] connections [in-person] to other methods”.

Other organizations identified that COVID-19 brought on an onslaught of barriers to their programming. Many organizations saw a complete end to the programs that they could offer. One community partner mentioned that “[they] really weren’t able to offer programming to the community”.

Overall, the impact of the pandemic on businesses largely varied, often a result of the nature of the organization itself.

### Improving the Functionality and Usability of the Systems Map

Upon discussing the usability and functionality of the systems map, community partners discussed a host of comments and suggestions in relation to the map (Figure 3). A majority of community leaders indicated that the creation of a systems map would prove useful and important for the Riverdale community.

**Fig 3.**
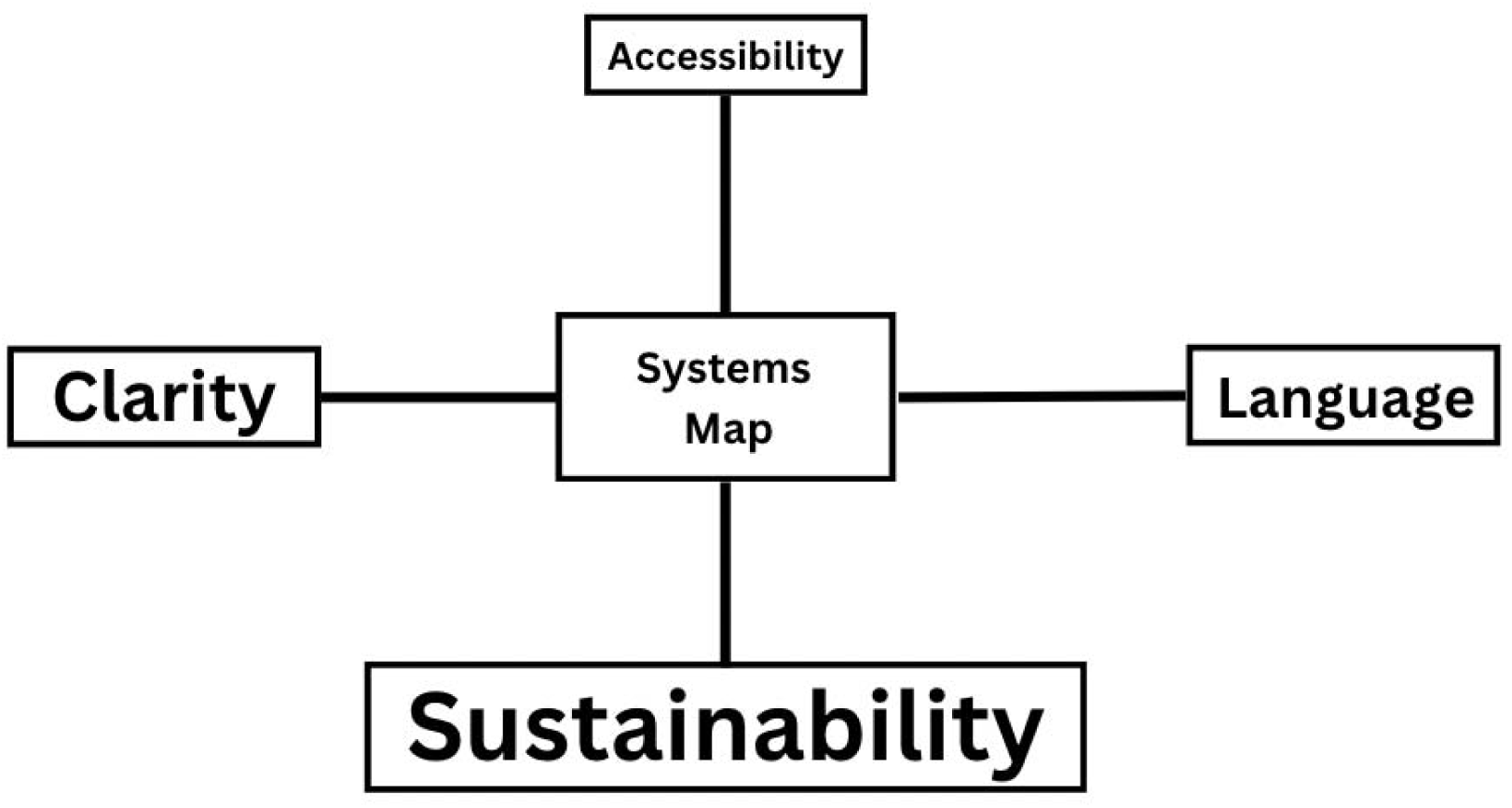
Themes associated with increasing usability and functionality of the systems map *Size of themes indicate their prevalence in analysis. Larger items indicate greater frequency in discussion.

#### Sustainability

Of all the discussion that occurred surrounding the systems map, sustainability of the tool was the biggest factor that would determine if the systems map would be both usable and functional. With regards to the map, whether information would be sustainable and accurate was in question.

> The biggest challenge with mapping is that as soon as they’re out of date, right as soon as they have information on them, that is just a little bit you know, not reliable, people don’t trust [the map anymore].

Participants also discussed how sustainability revolves around constantly updating information and ensuring that organizations know who to contact when information should change.

> And what the biggest barrier to this work is, is that community services change all the time. So, programs change all the time, organizations change all the time. They open they close, because it’s you know, a lot of these, a lot of this work is dependent on external funding through government or through different funding streams. So, I guess … this work is certainly needed and helpful because I feel like there isn’t really a transparency of what exists within our neighborhoods or what exists. You know, those things that keep us healthy and moving within our communities. But it’s just that piece around staying relevant. So, in five years from now, you know who will [manage] this? … So, I would just say like, I think that something like this is certainly helpful, but I guess there’s just like the sustainability piece of it.

Participants mentioned a few other resources that had attempted to accomplish the same mission, such as “211 Ontario” and “Red Pages”. However, the lack of sustainability of these resources is what ultimately led to their downfall.

#### Clarity

Another area of concern surrounding the usability and functionality of this tool was with regards to the clarity of the map itself. When showing individuals, some mentioned that it seemed “cluttered”. Individuals also desired the need for their services to be represented “a little bit clear[er]”. In fact, one community leader said:

> So I would always prefer something [with] less wording, less writing, in the maps. Something that has more ability to address everyone right … Yes, it might be helpful for certain people, but there might be [a need for] more simplified maps for families who are deal[ing] with linguistic barriers, also educational barriers.

Through iterative feedback from community leaders, adjustments to the map were made to “keep the information very simple and clear”.

#### Language

Some community leaders were eager to point out that Riverdale residents experience language barriers. Consequently, individuals addressed the need for language translations to be available on the systems map. One leader’s first question was “Can you translate this?”. Individuals also echoed that knowing one’s audience was a huge factor surrounding the functionality of this map.

> I would definitely encourage our youth to use this map. And I’m going to assume parents might predominantly use the map. So maybe considering the language barriers that might be there for … parents would be good … So, offering it in different languages, and using very simple terminology.

Using this piece of feedback, translations were made so individuals who speak Urdu, Pashto, and Arabic can easily access and understand the map.

#### Accessibility

Apart from language accessibility, accessibility when considering technological capabilities were also of concern. Knowing that the majority of individuals are from newcomer populations, comfort with using technology has to be considered. One community leader discussed the idea that “less clicks is better”. If an individual could easily navigate to the map, and navigate within it, that would be considered a success. One community leader also mentioned that inherently, this map is accessible, so folks do not have to “Google and [type] in whatever [they] have to type in to look at certain services”. This map provides a collective of resources that allows individuals to be aware of the local organizations that exist.

## Discussion

The perspectives of community leaders and services providers that are engaged in the delivery of healthy active living programs in Riverdale were explored in this qualitative descriptive study. The results of this study show the need for healthy active living programming, as well as describe the barriers and facilitators that are observed to impact service delivery including, 1) the ways in which programs are designed based on community needs, 2) the ways in which barriers and facilitators of program participation often directly parallel one another, and 3) the way in which usability and functionality often require putting community needs at the forefront of intervention design and programming.

Based on interviews with program leaders, programming is always developed with the community’s needs at the forefront. It is developed so that individuals who are within the population are always served. Based on the demographics and needs of the Riverdale community, community partners have catered programs specific to certain demographics. Whether considering age, gender, or cultural background, community programming is designed to meet and aid community’s needs. An understanding of these factors is almost always determined by organizations, many of which include members of staff who have similar experiences to those of the Riverdale community. However, some organizations do not possess these personal connections. Consequently, they often consult and check in with the needs of their client base. To reach the newcomer populations of the Riverdale community, many organizations hire individuals who speak multiple languages, so that these individuals can host and run programming, or translate as needed. Apart from a general understanding of the types of users that engage in programming, it became clear that there was a lack of representation for those in the LGBT2SQIA+ community.

Separate from understanding the needs of the community, and using that to develop programming, community leaders are also hyper-aware of the barriers and facilitators that exist within the populations that they serve. Through understanding the needs of their communities, program leaders have taken it upon themselves to understand the reasons behind the amount of engagement in their programs. Program leaders are deeply tied to their communities, and consequently, have a very good understanding of the types of barriers that exist. In an effort to provide support and programming to Riverdale newcomers, these leaders have developed ways of mitigating these barriers. Often, this involves directly combatting specific barriers. For instance, through recognizing that financial barriers exist, community leaders have created programming that can help to alleviate this stress. In fact, provision of food, provision of transportation methods, and even compensation are some examples of how program staff are trying to help and support those in the Riverdale community. Although this is a step many community leaders would like to build upon, by addressing more barriers, funding often poses an issue. Community leaders are often faced with the issue of funding programs, with few participants. Or conversely, programs are funded, but they are not funded enough to ensure that everyone who wants to, can participate. Apart from barrier and facilitator discussion, it became clear through the interviews, that there is not a central resource that individuals can access to gain an understanding of local programming. In fact, it often happens through word-of-mouth – highlighting the need for a tool like the systems map.

Upon review of the systems map, it became clear that a majority of community leaders thought it would be a useful tool. Not only would it provide the Riverdale community with a platform where they can learn about what is offered, but it would allow the community organizations to accurately show and represent their programming. With the enthusiasm surrounding the systems map, there were a few caveats. Individuals expressed their concern about continuity and sustainability of this tool. With many community leaders being newcomers themselves, there was a general agreement that the map should be accessible from a technological and language standpoint. Using the feedback provided from these interviews, iterative changes have been made to the map.

### Future Directions

From the findings of this study, the next steps have been revealed. First, understanding the types of programming that are available, and the users that are engaged in such programming has revealed that more work must be done to create open and positive spaces for those in the LGBT2SQIA+ communities. In addition to this, working to promote programming through the use of the systems map is something that can benefit many community organizations. Doing so, however, requires the systems map to be sustainable, clear, and accessible. To accomplish this, iterative discussions will need to be had within the SCORE! team to discuss the means in which to sustain this resource. Further, feedback from the community and desired users of this tool would be beneficial in ensuring the map remains clear and accessible to the target population. Although adjustments have been made to the map to ensure that some accessibility and clarity needs are met, the map will require constant review and editing to ensure that information is as accurate as possible. Consequently, the future of this map, and the plan to maintain this map after the completion of the SCORE! project must be considered and discussed.

### Conclusion

This qualitative research study was done with the intent of understanding the perceptions and observations of service providers and community leaders in the Riverdale neighbourhood on local programs, the barriers and facilitators of said engagement, and the functionality of a systems map. Through this work, it is clear that the Riverdale community has many programs that meet the needs of many. Further, the creation of a systems map may be beneficial in promoting program participation and awareness. Through this recognition, continual and sustained steps must be taken to increase healthy active living programming and their relative newcomer participation.

It is recognized that the systems map will miss some of the some of the services. However, it provides a summary of the services in the Riverdale neighbourhood that can be updated as new information is revealed. It is also recognized that the service landscape will change over time. Once again, the systems map can be updated as the environment changes.

## Data Availability

All data produced in the present study are available upon reasonable request to the authors

## Declarations

### Ethics Approval

This study was approved by the Hamilton Integrated Research Ethics Board (HIREB #15120). Informed consent was obtained from all individual participants included in the study.

### Consent to participate

Participants were asked to provide informed consent upon participating.

## Appendix A: Interview Guide

1. Could you please share the following details about your organization?

- Organization’s size
- Mission, vision, values
- Do you offer any services in any languages other than English?
- Key target demographics (i.e., families, youth, elderly, etc.)
2. What activities are utilized most at your organization/program?
3. Who engages most with said program/activity?

- Prompt: Are there mainly boys or girls participating?
- Prompt: What is the age range of each group?
4. Have you noticed any gender-based differences amongst people that visit the organization?

- Do more girls show up, or boys?
5. Have you ever had transgender, non-binary, or gender non-conforming participants?

- Yes – Is there any feedback from them or observation from you that you could share?
6. What are the peak hours experienced by your organization?

- Prompt: Do you notice any changes according to the season? Do peak hours change in summer or winter?
7. Are you aware of any barriers to accessing your services experienced by users?

- Prompt: Has the COVID-19 pandemic negatively affected people’s access to services or reduced participation?
8. Are you aware of any facilitators to accessing your services experienced by users?

- Prompt: Have you ever had promotional offers, or offered transportation/carpool offered?
9. Are your services/programs accurately represented on this map?

- Prompt: If not – What should change?
10. Are there any additional/related programs and services you think should be included in this map?
11. Do you think this map would be useful to Riverdale residents?
12. Do you know the ethnicity or race of people who attend these events/activities?
13. Do you get families/interested clients who may have language barriers?
14. How do the activities at your organization/program interact with existing resources in the community?
15. What things encourage people to stay engaged in activities at your organization/program?
16. Is there anyone else you think we should interview?

